# Sars-Cov-2 in Argentina: Following Virus Spreading Using Granger Causality

**DOI:** 10.1101/2020.10.06.20207993

**Authors:** Juan M.C. Larrosa

## Abstract

There is a debate in Argentina on how COVID-19 outbreak in one district ends up infecting its neighbor districts. This contribution aims to use tools of time series analysis for understanding processes of contagious through regions. I use VAR and Granger causality for testing neighbor spreading via sequential rate of contagion. Results show that in the case of Argentina, contagion began in the capital city of Buenos Aires and then spread to its hinterland via specific districts. Once interior districts were infected a positive feedback dynamics emerge creating regions of high reproducibility of the virus where interventions may be focus in the very near future. This specific use of time series analysis may provide a tool for tracing infectiousness along regions that may help to anticipate infection and then for intervening for reducing the problems derived by the disease.

## 1. Introduction

The purposes of this contribution is to econometrically analyze the sequences of contagious and tests with the mobility of the population. In a national embarrassing series of political discussions on the isolation measures adopted by the central government and other political divisions of the country, diverse topics have emerged with crossed accusations among diverse national actors. First, there were unbelievable discussions on whether the contagion initiated in one territory has purposively derived to a neighbor district[1]. Second, there were also discussions on whether relaxation of capital city strict isolation restrictions adopted by the central government may increase the rate of contagion of districts distant but connected to the former [2].

In a pandemic case, outbreaks are geographically located. Once contagion begins infected individuals may move through another geographical points spreading the infection. Assuming everything else equals, contagion rate at one point in time in one place could be associated to a posterior contagion rate in the vicinity if mobility, even reduced by lockdown, is allowed. I follow the rate of contagion con COVID-19 in the most populated area of Argentina, the Buenos Aires Metropolitan Area (BAMA) that accounts for approximately 16.5 million inhabitants. The area comprehends the capital city (Buenos Aires Autonomous City or BAAC) and the surrounding political divisions called parties (*partidos*) or departments that belong to another district: the Buenos Aires province (BAP). Nearest parties to BAAC are called the First Conurbation Cordon (FCC) and the following ring is called Second Conurbation Cordon (SCC). Usually FCC and SCC provide a daily flow of workers for the firms and state entities arranged in BAAC. There is a natural connection and sequence of rate of contagion among these three regions. As absurd as it might seems, national and Buenos Aires province authorities made repetitive and direct accusations to the head of BAAC (governed by the political opposition) on relaxing controls that made possible that infected people spread the virus from to capital to the province.

Following the sequence of the rates of contagion of each of these districts time series tools can be used for detecting causal inference and shocks from one time series to the other. Did Buenos Aires city infected people spread the virus in the nearer Greater Buenos Aires and how? What other pattern of contagion is inferred from data analysis? I use data on registered confirmed cases in each district and then follow its path by estimating Granger causality and impulse-response functions from a VAR modeling approach. In the best of our knowledge few reports have studied the Argentinian case in terms of contagion data (an exception is [3]). I find sequences of rate of contagion of districts influencing the rate of contagions of neighbors and then creating patterns of mutual contagion between specific districts.

## 2. Spreading from BAAC to BAP: Modeling approach

I model at first the statement that contagion generated in BAAC (*y*_BAAC_) then spread to the BAP. Specifically, the nearest BAP districts to BAAC. The FCC is composed by Avellaneda (*y*_*fa*_), Gral. San Martín (*y*_*fsm*_), La Matanza (*y*_*flm*_), Lanús (*y*_*fl*_), Lomas de Zamora (*y*_*flz*_), Morón (*y*_*fm*_), San Isidro (*y*_*fsi*_), Tres de Febrero (*y*_*ftf*_), and Vicente López (*y*_*fvl*_) parties. The SCC, on the other side, is composed by Almirante Brown (*y*_*sab*_), Berazategui (*y*_*sb*_), Escobar (*y*_*se*_), Esteban Echeverría (*y*_*see*_), Ezeiza (*y*_*se*_), Florencio Varela (*y*_*sfv*_), Hurlingham (*y*_*sh*_), Ituzaingó (*y*_*si*_), José C. Paz (*y*_*sjcp*_), Malvinas Argentinas (*y*_*sma*_), Merlo (*y*_*sme*_), Moreno (*y*_*smo*_), Pilar (*y*_*sp*_), Quilmes (*y*_*sq*_), San Fernando (*y*_*ssf*_), San Miguel (*y*_*ssm*_), and Tigre (*y*_*st*_) parties. Figure 1 displays the respective map. BAAC is painted in orange, the FCC is in light grey and the SCC is painted in dark grey.

**Figure 1.**
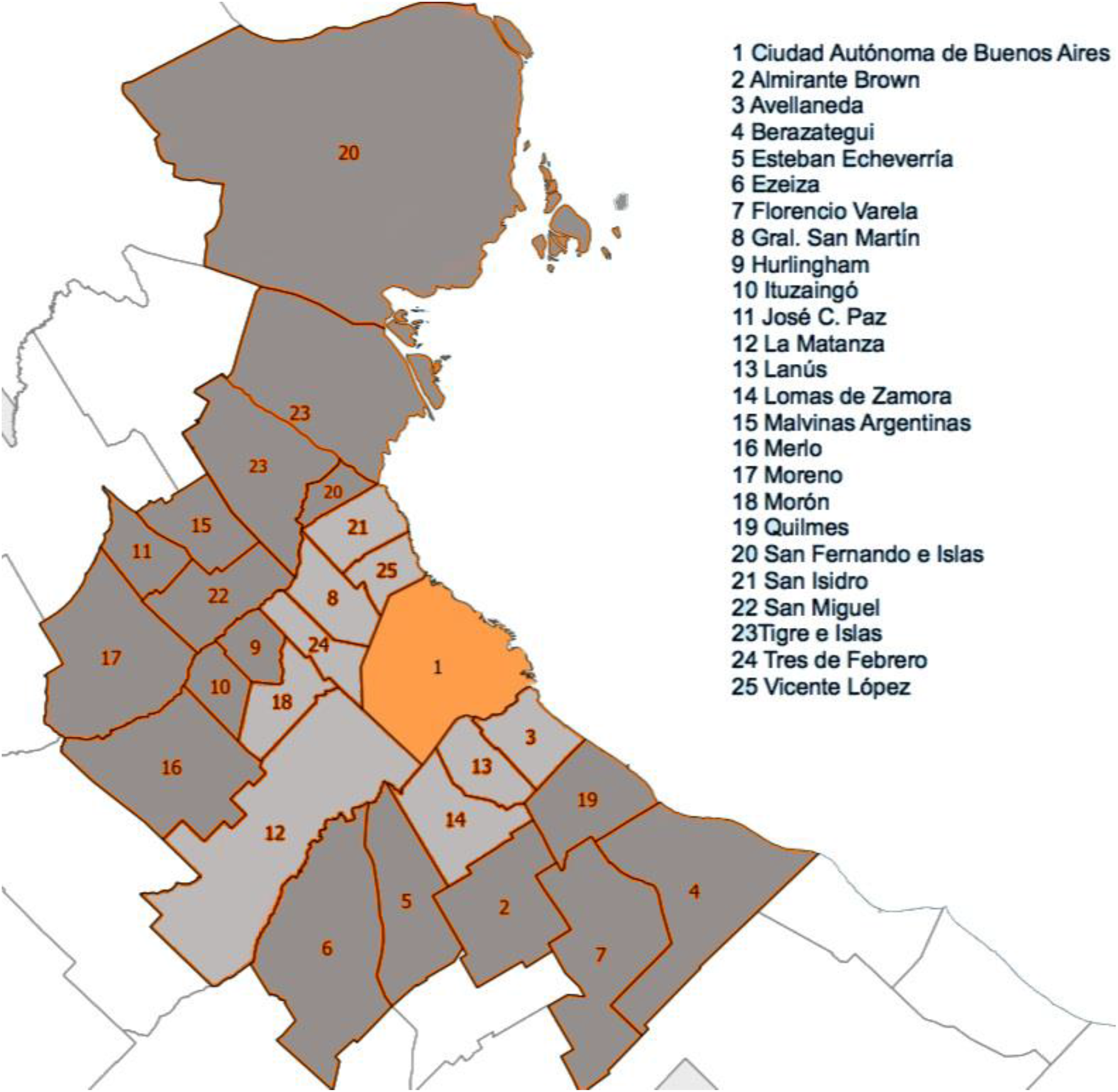
BAAC (Ciudad Autónoma de Buenos Aires), FCC (light grey), and SCC (dark grey)

By having the rate of contagion on daily basis of the specific parties of each cordon it can be asked questions to the data by modeling these rates of contagion as interrelated. I rely on basic vector autoregressive representation such as described next. VAR representation for COVID-19 studies has shown that for international comparisons represents an interesting tool for answering short-run dynamics in contagion [4].

### 2.1 Vector Autoregressions and Granger Causality

Assume the *n*-variable vector time series *y*_*i*,*t*_ evolves according to the following model:

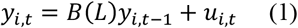

where *B*(*L*)*y*_*i*,*t*−1_ = *B*_1_*y*_*i*,*t*−1_ *+ B*_2_*y*_*i*,*t*-2_ *+ …*.*+ B*_*p*_*y*_*i*,*t*-*p*_ and u_*t*_ is normally distributed rand error vector with zero mean and covariance matrix Σ. I then define a (1) as a *p*^*th*^ –order VAR or VAR(*p*) given the number of *p* lags included. It also may be included time trends, constants or seasonal dummy variables. Assuming stationarity in the VAR it can be denoted a moving average representation of (1) as

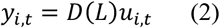

with *D*(*L*)u_*i*,*t*_ = (*I - B*(*L*))^−1^. In the moving average representation *y*_*i*,*t*_ is written as a linear combination of u_*i*,*t*_. This moving average representation lies in the basis of computing the impulse-response functions and error variance decompositions. On the other hand, a reduced-form VAR results a plausible way of observing second-moment properties of a group of time series. This representation is usually applied for testing Granger-causality inferred when a time series and its lags are statistically significant in explaining other/s time series.

Impulse-response functions (IRF) and forecast error variance decompositions (FEVD) are the sub products of VAR estimation and an ideal approach for studying second-moment properties based on (2) portraying how predicted values of the variables of the model are influenced by the error terms at different time spans. IRF will present a graphical display on time (steps) showing how one shock (one standard deviation) on one variable will affect the other variable. The research will focus on how this initial shock affects by observing if this initial effect may be null, or have a positive or negative effect and if this effect tends to converge in time to zero and disappearing in time or if the shock tend to have permanent effects on the affected variable. See [5] for an in-depth of VAR modeling.

The statistical exercise is as follows. I assume contagion in one district may spread contagion in its neighbor districts. Data was obtained on district rate of contagion in [6]. I model each rate of contagion as dependent of the rate of contagion of its neighbor districts. I model these interrelations through a VAR model of rate of contagion, where lags are selected to 14 given that on average symptoms emerge only as earlier as 5-6 days, and at most up to 14 days. This way it contemplates the incubation period estimated by the virus since infection to first symptoms [7]. Then it is measured the directionality of the contagion by Granger causality and the intensity of this process by observing specific impulse-response functions from VAR system. As I only consider interactions among neighbor parties calculations were reduced from 600 (24 × 25 districts) to 77 in the case of Granger causality as a result.

These assumptions may have its weaknesses. Firstly, parties are highly interconnected given the dense urban structure and highways and train infrastructure that easily mobilizes people from one party to another non-neighbor district. Secondly, I do not control for health infrastructure differences that surely plays a role. Thirdly, contagion is a complex process simplifying just by closeness in our case.

## 3. From BAAC to FCC and then to SCC

As BAAC registered the first cases by travelers arriving from infected zones overseas [8], a priori it is known the origin and from that point on it can be inferred the sequence of contagions where it spreads. Figure 2 depicts the daily number of confirmed cases in the BAAC, its neighboring FCC, and the outer ring of these two, the SCC. As observed, BAAC surpassed the two regions initially but by early June these two largely surpassed the former in increasing numbers. It is also observed that BAAC reaches a plateau by mid-July. This way, a question of whether there were causality of contagions remain unanswered.

**Figure 2.**
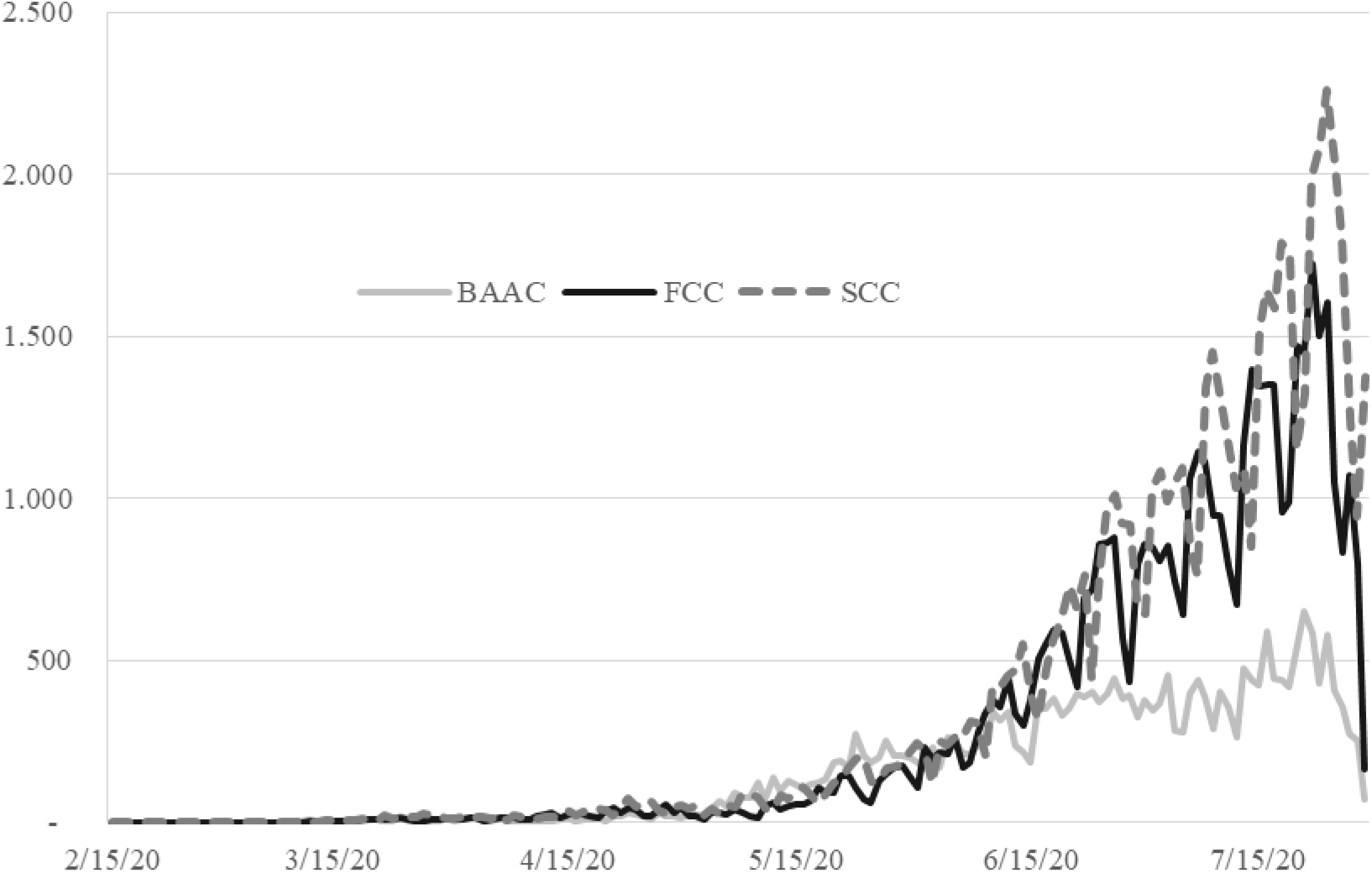
Rate of Daily Confirmed Cases by Regions.

**Figure 3.**
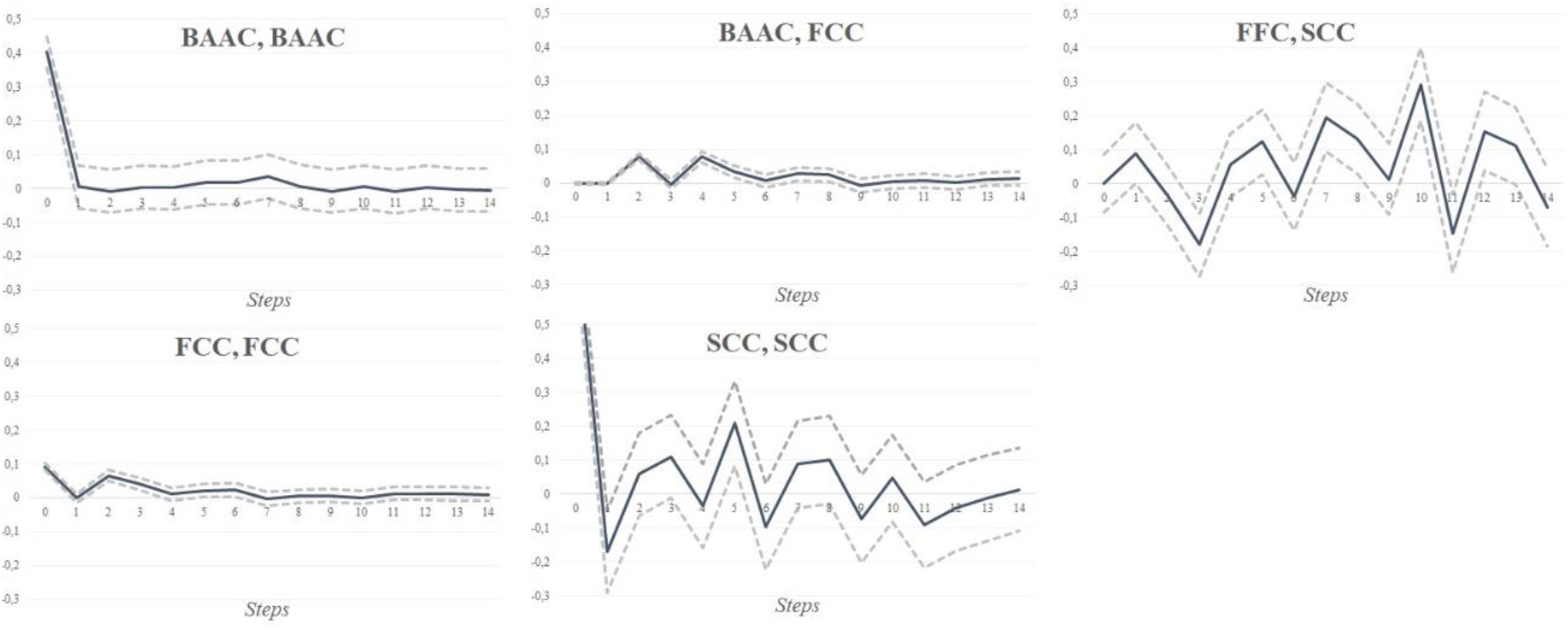
Orthogonalized Impulse-Response Functions (95% Confidence Interval) BAAC to FCC and SCC.

Just to shed light to this matter, it is performed a simple Granger causality [8] exercise with the rate of contagion (confirmed cases) of the three regions with daily data from February 15^th^ to July, 29^th^. The rate of contagion of the three regions are stationaries granted statistical interpretation to the results. *y*_BAAC_, *y*_FCC_, and *y*_SCC_ are the name of these variables respectively. I estimate a simple VAR and then tested for Granger causality. As prior procedure it has been tested that the time series are stationary according Dickey-Fuller unit root test for more than 14 lags and that lag structure is used as explained earlier. Table 1 shows the results: the rate of contagion of BAAC does not Granger-cause in terms the rate of contagion of FCC nor the SCC. The rate of contagion of the FCC is significantly Granger caused by BAAC and in a lesser significance by the SCC. Finally, the rate of contagion of BAAC and FCC cause in terms of Granger the rate of SCC.

**Table 1.** Granger Causality on BAAC to FCC and SCC.

| Equation | $\chi^2$ | df | Prob > $\chi^2$ | Summary |
| --- | --- | --- | --- | --- |
| $y_{BAAC} \leftarrow y_{FCC}$ | 1.6274 | 14 | 1.000 | Does not Granger cause |
| $y_{BAAC} \leftarrow y_{SCC}$ | .41253 | 14 | 1.000 | Does not Granger cause |
| $y_{BAAC} \leftarrow \text{All}$ | 2.2811 | 28 | 1.000 | Does not Granger cause |
| $y_{FCC} \leftarrow y_{BAAC}$ | 4857.5 | 14 | 0.000 | Does Granger cause |
| $y_{FCC} \leftarrow y_{SCC}$ | 31.041 | 14 | 0.005 | Does Granger cause |
| $y_{FCC} \leftarrow \text{All}$ | 5257.8 | 28 | 0.000 | Does Granger cause |
| $y_{SCC} \leftarrow y_{BAAC}$ | 95.132 | 14 | 0.000 | Does Granger cause |
| $y_{SCC} \leftarrow y_{FCC}$ | 97.475 | 14 | 0.000 | Does Granger cause |
| $y_{SCC} \leftarrow \text{All}$ | 339.02 | 28 | 0.000 | Does Granger cause |

Finally, the impulse-response functions of the VAR of the three variables indicate that a shock of one standard deviation in the rate of contagion ends up affecting strongly the SCC. First, a shock on BAAC rate of contagion from its own rate affects it strongly one day and then dissipates. In the same case for FCC is less shocking and then it also tends to dissipate. A shock from BAAC to FFC has effects in 2-4 days and then also dissipates. However, a shock from FCC to SCC alter the rate of contagion of the later and its effect remains strongly during the period. It shows the same effect if the shock come from the same region. This would mean that SCC is highly sensitive to outside and internal shocks and once it receives the impact it does not dissipate in time and that keep increasing the rate of contagion.

First answer. The rate of contagion of BAAC indeed Granger-causes the rate of contagion of the neighboring conurbation cordons but the more affected region is SCC that received immediate shocks from FCC and more distant shocks from BAAC (most probably repercussions of BAAC affecting FCC and then cascading to SCC).

## 4. A Path of Contagion

Evidence has been presented of cascading from BAAC to FCC and then to SCC and that SCC once receives the shock to the rate of contagion it persists. Now it can be dissagregated the problem by observing where was the paht of contagion from the BAAC to the SCC by repeating the procedure considering now the rate of contagion of each party related to the rate of contagion of its neighbors.

Table 2 presents the Granger causality on rates of contagion from all 25 limiting districts. Most of the causality relations are bi-directional in terms one rate causing the other and viceversa. Interestingly, BAAC (*y*_BAAC_) and La Matanza (*y*_*flm*_) are not affected by contagion in most of neighbors but Granger-causes the rate of contagion of them. This way, it seems that the main path of contagion initiates in BAAC and goes to La Matanza and once there, as seen previously, and reinforced by Table 2 information, it has spread it in almost all directions.

**Table 2.**
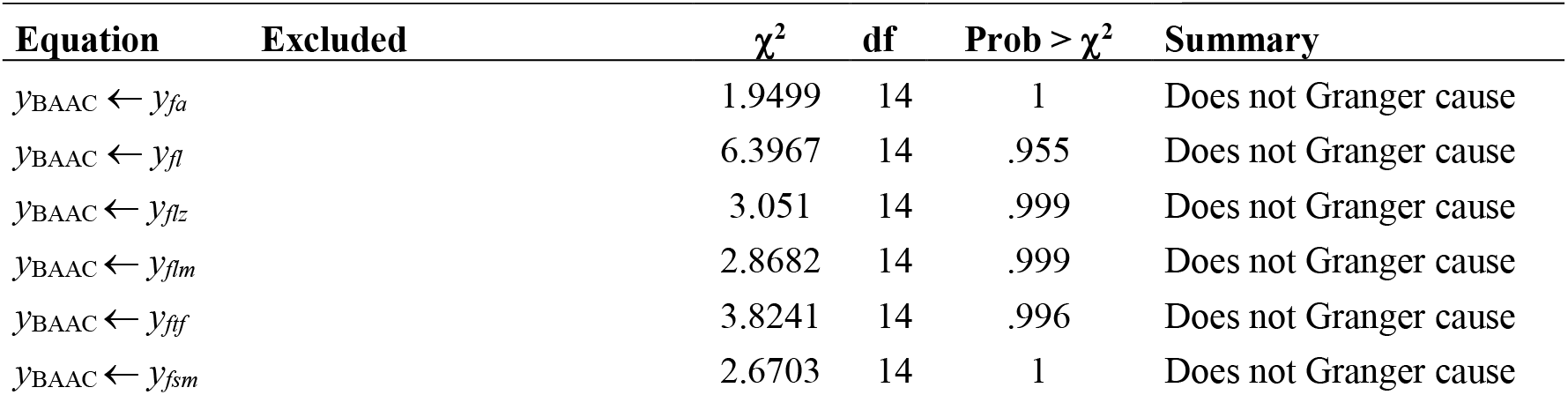
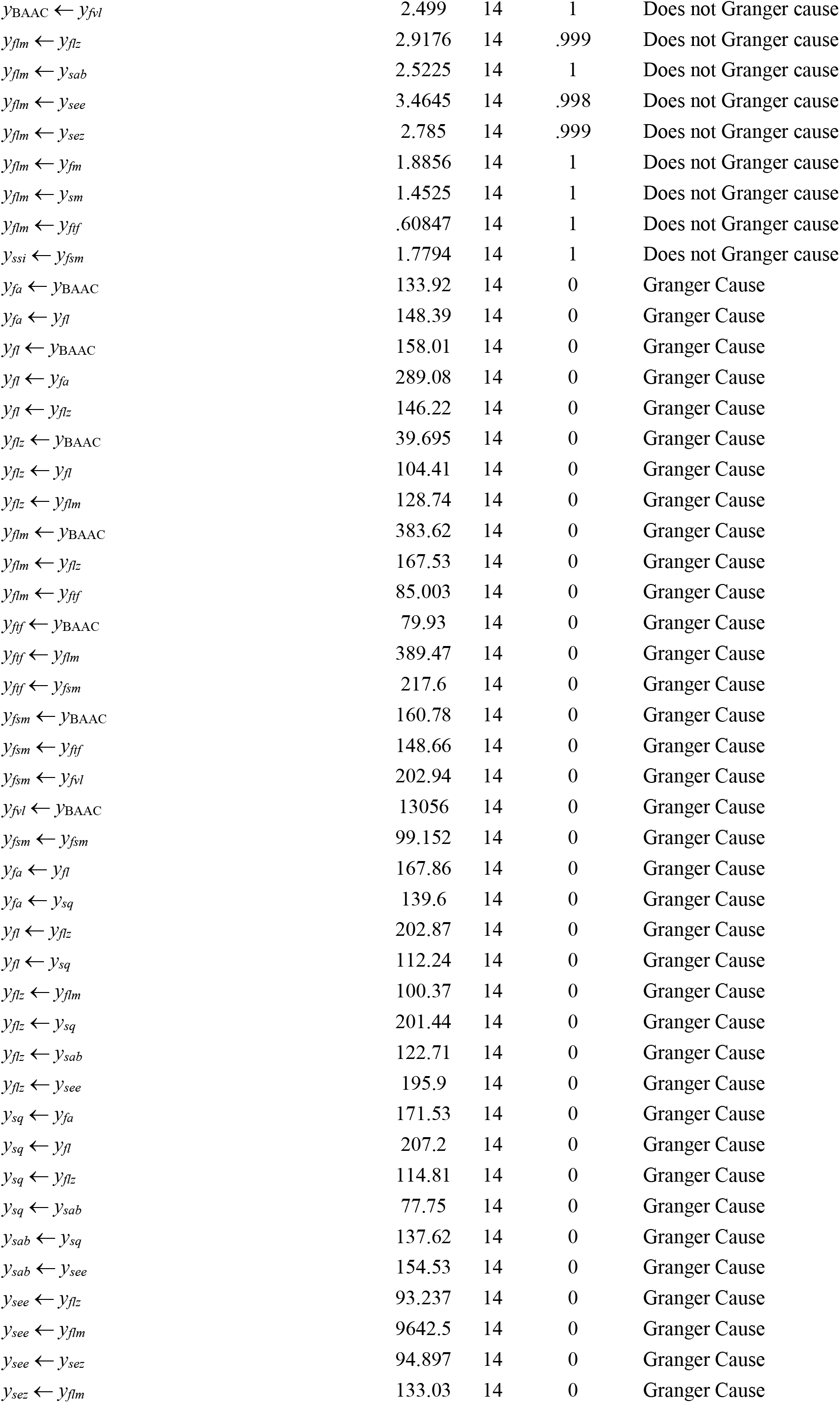
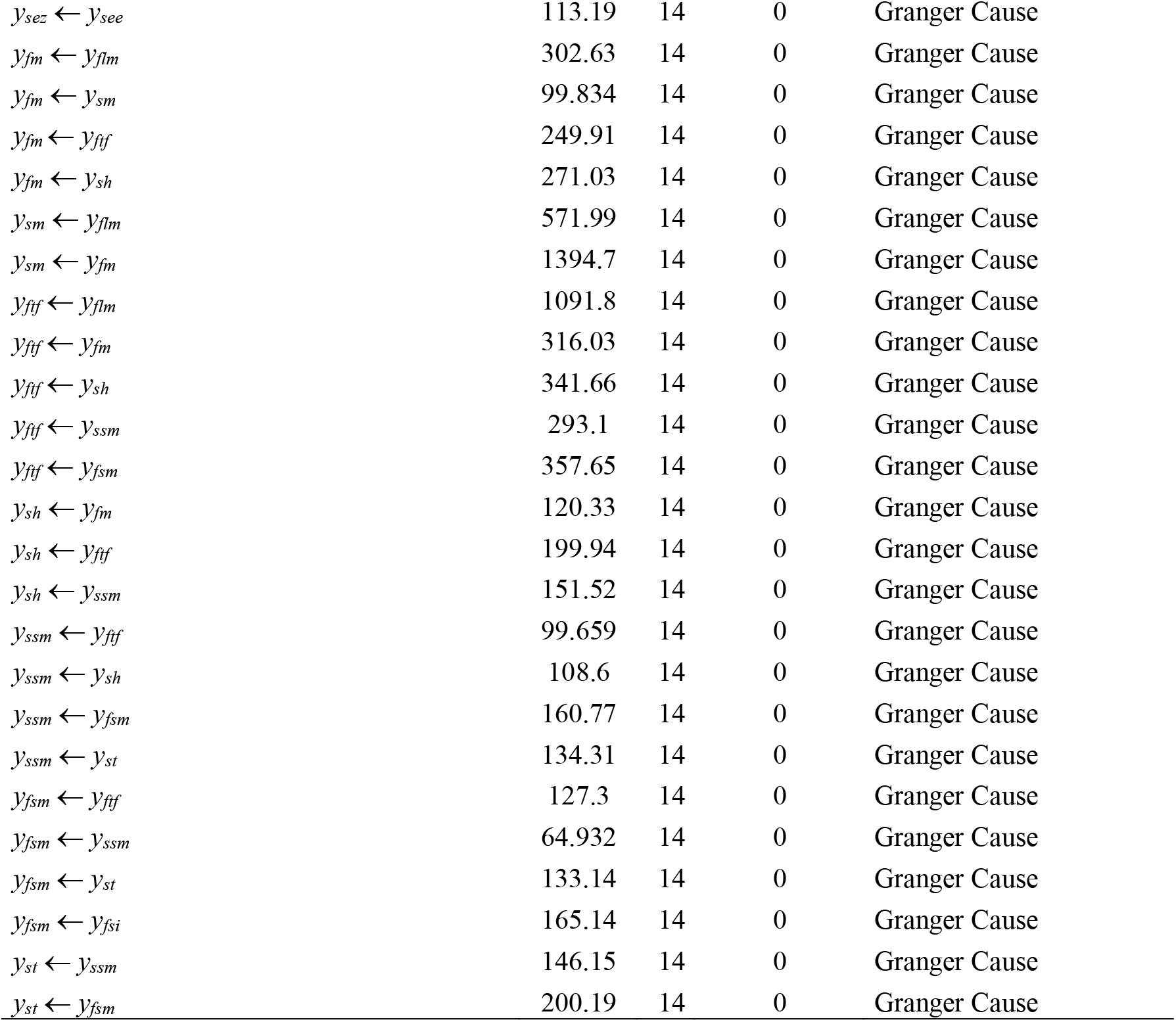
Granger causality of rate of contagion among limiting parties.

In terms of intensity Figure 4 presents the most significant IRF selected from the analysis of the complete set of estimations. It is not displayed the majority of IRF given that shocks affected in a negligible way the affected rate of contagion (usually impact and following steps of less than .01). This selection shows other potential paths of contagion from BAAC to Vicente López and its neighbors (at the North of BAAC). Then from the South, BAAC shocks to Lanús and La Matanza seems to take a week or two to affect the respective rates of contagion. Following the path leads to a triangle of shocks affecting mutually the Berazategui, Moreno, and Quilmes parties and on the other corner it can be observed sensitive mutual shocks among Moreno, Ituzaingó, and Merlo.

**Figure 4.**
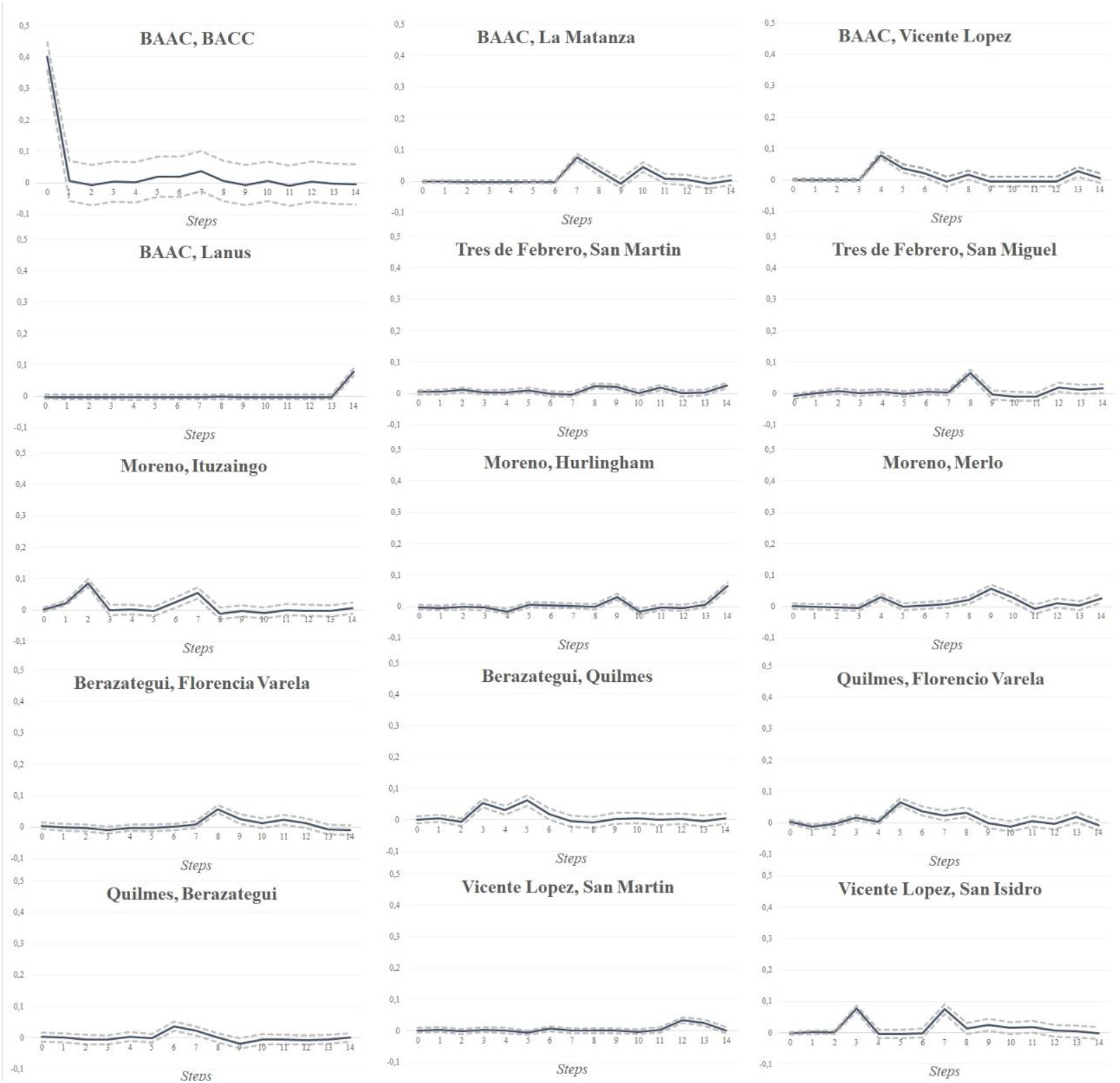
Main Orthogonalized Impulse-Response Functions (95% Confidence Interval) by Parties.

The information on directionality and intensity can be represented in a network of districts as sketched in Figure 5. Green edges (lines) represent bidirectional Granger causality and red ones represent one-way causality. The size of node is proportional to the natural logarithm of infected cases. The thickness of edges illustrates specific scales in IRF from rate of contagion of one party to the other. Contagion began from BAAC as reported and the spread via Vicente López to the North and via La Matanza to the South. Once in the SCC it creates feedback of contagion between selected parties in the South (Berazategui, Quilmes, and Florencio Varela) and the West (Moreno, Merlo, Hurlingham). These last are precisely located at the SCC that helps to explained the non-convergent dynamics detected previously.

**Figure 5.**
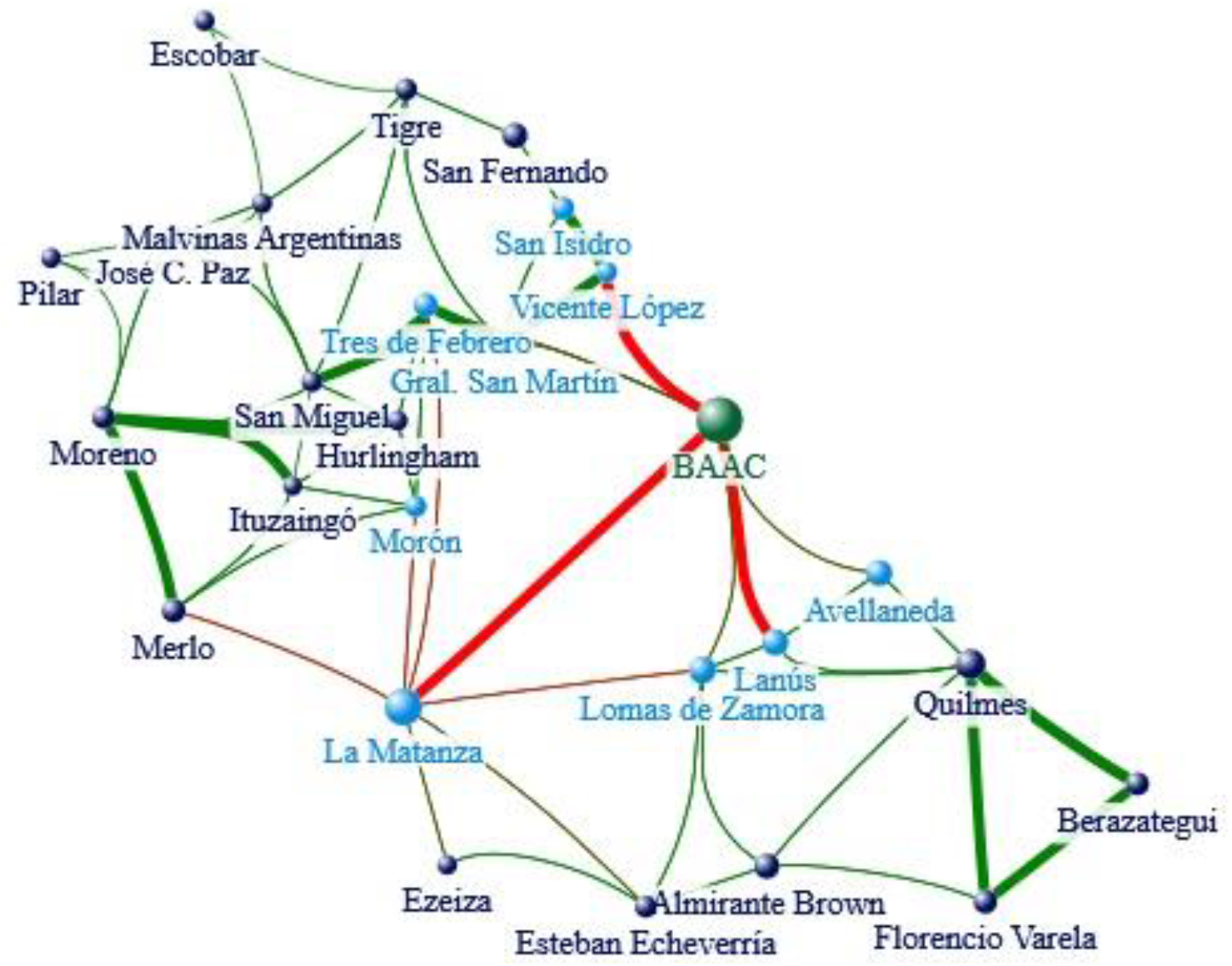
Granger Causality (direction) and Significant Impulse-Response Cases (intensity) in the BAMA.

By tracing the contagion it is feasible to respond to different questions specially in terms of spreading time, available infrastructure of the target places, and detection of mutually contagious regions where the virus (in our case the COVID-19 but it could be any other contagious menace) circulates feeding back other contagion accelerating them exponentially. These are regions of urgent intervention.

## 5. Conclusions

I use Granger causality and IRF to understand how contagion emerges in one region and then sequentially spreads to its hinterland. I use simple data on rate on contagion publicly available and detects clues on a rate of contagion initiating an outbreak and then correlating in time with contagions in neighbor interconnected regions. I disaggregate the information and trace district by district the contagion and reveals dynamic of contagions where regions infect their neighbors but are not being infected by them demarking a path of contagion. Once revealed it specific dynamics are identified between districts that would deserve more attention because of the presence of positive loops of contagion.

The present work can be easily reproduced in many countries under the COVID-19 pandemic and it would serve as a tracing technique in the absence of more concrete microdata of infected people, their comorbidity data, and contact chains.

## Data Availability

All data is available at DOI: 10.13140/RG.2.2.16398.18244

https://doi.org/10.13140/RG.2.2.16398.18244

## Appendix

## References

[1] BA Times, “Buenos Aires versus Buenos Aires”, 05-23-2020 [https://www.batimes.com.ar/news/opinion-and-analysis/buenos-aires-versus-buenos-aires.phtml]

[2] BA Times, “Nerves fray in Argentina, as some flout pandemic lockdown”, 08-13-2020 [https://www.batimes.com.ar/news/argentina/nerves-fray-in-argentina-as-some-flout-pandemic-lockdown.phtml]

[3] Ahumada, H., Espina, S. y Navajas, F. (2020), COVID-19 with Uncertain Phases: Estimation Issues with An Illustration for Argentina (June 20). Available at SSRN: https://ssrn.com/abstract=3633500 or http://dx.doi.org/10.2139/ssrn.3633500

[4] Haffner, C.H. (2020). The Spread of the Covid-19 Pandemic in Time and Space. Int. J. Environ. Res. Public Health 2020, 17(11), 3827; https://doi.org/10.3390/ijerph17113827

[5] Amisano, Gianni and Carlo Giannini (1997), Topics in Structural VAR Econometrics. Springer Verlag,

[6] Ministry of Health, COVID-19. Casos registrados en la República Argentina, Downloaded 7/29/2020 at http://datos.salud.gob.ar/dataset/covid-19-casos-registrados-en-la-republica-argentina

[7] Stephen A. Lauer, Kyra H. Grantz, Qifang Bi, Forrest K. Jones, Qulu Zheng, Hannah R. Meredith, Andrew S. Azman, Nicholas G. Reich, and Justin Lessler (2020) “The Incubation Period of Coronavirus Disease 2019 (COVID-19) From Publicly Reported Confirmed Cases: Estimation and Application”, Annals of Internal Medicine, https://doi.org/10.7326/M20-0504

[8] BA Times (2020), “Health Ministry confirms first cases of ‘community transmission’ in Argentina”. [https://www.batimes.com.ar/news/argentina/health-ministry-confirms-first-cases-of-community-transmission.phtml]

